# Two-Month Study on Acute Poisoning Cases in a Tertiary Care Hospital in Bangladesh

**DOI:** 10.1101/2025.03.21.25324397

**Authors:** Sabbiha Nadia Majumder, Md. Robed Amin

## Abstract

**Introduction:** This study presents an observational analysis of acute poisoning cases admitted to Dhaka Medical College Hospital, Bangladesh, over a two-month period, highlighting key demographic, etiological, clinical, and outcome-based trends.

**Results:** A total of 611 cases were reviewed, with a predominant incidence among young adults aged 20-40 years. The highest fatality rate occurred in the 16-25-year-old cohort, with a secondary fatality peak noted among individuals aged 66-70, indicating varying vulnerability across age groups. Pesticide-related poisonings were common, with nearly equal distributions between organophosphorus compounds (51%) and non-organophosphorus compounds (49%). Commuter poisoning (31.2%) was notably the leading type of non-pesticide poisoning, primarily associated with criminal activities. Suicidal poisoning accounted for most cases at 57.4%, often linked to family disharmony as a frequent underlying factor. Additionally, snake bites (7.5%) emerged as a notable public health concern, especially in rural areas.

Fatalities were highest among those who ingested kerosene (20%), despite its low incidence. Logistic regression analysis identified key predictors of mortality, including severely low Glasgow Coma Scale scores (GCS<9; OR=80.59, p<0.001), hypotension, tachycardia, and pupillary abnormalities. Non-organophosphorus poisonings displayed increased severity in terms of neurological impairment and hypotension compared to organophosphorus poisonings, which mainly resulted in pupillary constriction.

Demographic analysis revealed a higher mortality risk among females, rural residents, and individuals with lower educational attainment, although the statistical significance was limited.

**Conclusion:** This study identifies important gaps in managing and preventing acute poisoning in Bangladesh. It calls for structured clinical protocols, better training for healthcare providers, and targeted community interventions. Improving public awareness and timely medical responses is essential to reduce poisoning-related morbidity and mortality.

## Introduction

Acute poisoning is a significant cause of visits to emergency departments and hospital admissions in Bangladesh, leading to substantial morbidity and mortality.^1^ Despite being largely preventable, managing poisoning cases poses challenges due to gaps in identification, clinical evaluation, and the availability of evidence-based protocols.

This study aims to examine trends and provide data to enhance preparedness and response capabilities for acute poisoning. This observational study explores the demographic profile, nature, aetiology, and short-term outcomes of acute poisoning cases admitted over two months at Dhaka Medical College Hospital in Bangladesh. Acute poisoning has a considerable impact on healthcare resources and patient outcomes. Therefore, this research aims to highlight critical areas that require intervention and improve the healthcare system’s readiness.

## Objective

The study aimed to categorize poisoning cases, identify causative agents, assess demographic factors, determine modes of poisoning, and evaluate short-term outcomes of these cases in a tertiary hospital.

## Methods

A cross-sectional observational study was conducted in all adult medicine units at Dhaka Medical College Hospital over a two-month period, from September 15 to November 15, 2017. The study included patients aged 14 years and older who presented with clinical features and a confirmed history suggestive of poisoning.

Cases were identified through relevant histories provided by patients and their attendants, as well as clinical toxidrome presentations.

Patients were excluded if they had ambiguous histories, food poisoning, non-poisoning-related unconsciousness, or if they refused to consent to the study.

Structured questionnaires were used to systematically record demographic data, the type and amount of poison ingested, the intention behind the poisoning, clinical signs, symptoms, and social backgrounds. Trained research associates conducted direct clinical observations to ensure accurate and detailed documentation of each case. Whenever possible, samples of the ingested poisons were collected from patients or their attendants for cross-verification. Additional information was gathered from hospital records, including nursing logs and treatment registers. Special attention was given to snakebite cases, with thorough descriptions of the snakes involved, supported by specimens provided by patients when available. Ethical approval was obtained from the Ethical Review Committee of Dhaka Medical College Hospital, ensuring compliance with ethical standards in medical research. Written informed consent was acquired from patients or their legal guardians. Statistical analysis was performed using SPSS version 20. Descriptive statistics, such as means, medians, frequencies, and percentages, were used to summarize demographic and clinical variables. Logistic regression analysis was conducted to identify factors significantly associated with mortality outcomes.

## Results

During the study period, 611 cases of poisoning were evaluated, although data for 39 patients were missing. The demographic analysis revealed variability in age distribution, with a notable predominance among younger adults (ages 20-40 years). (Fig 1)

**Figure 1:**
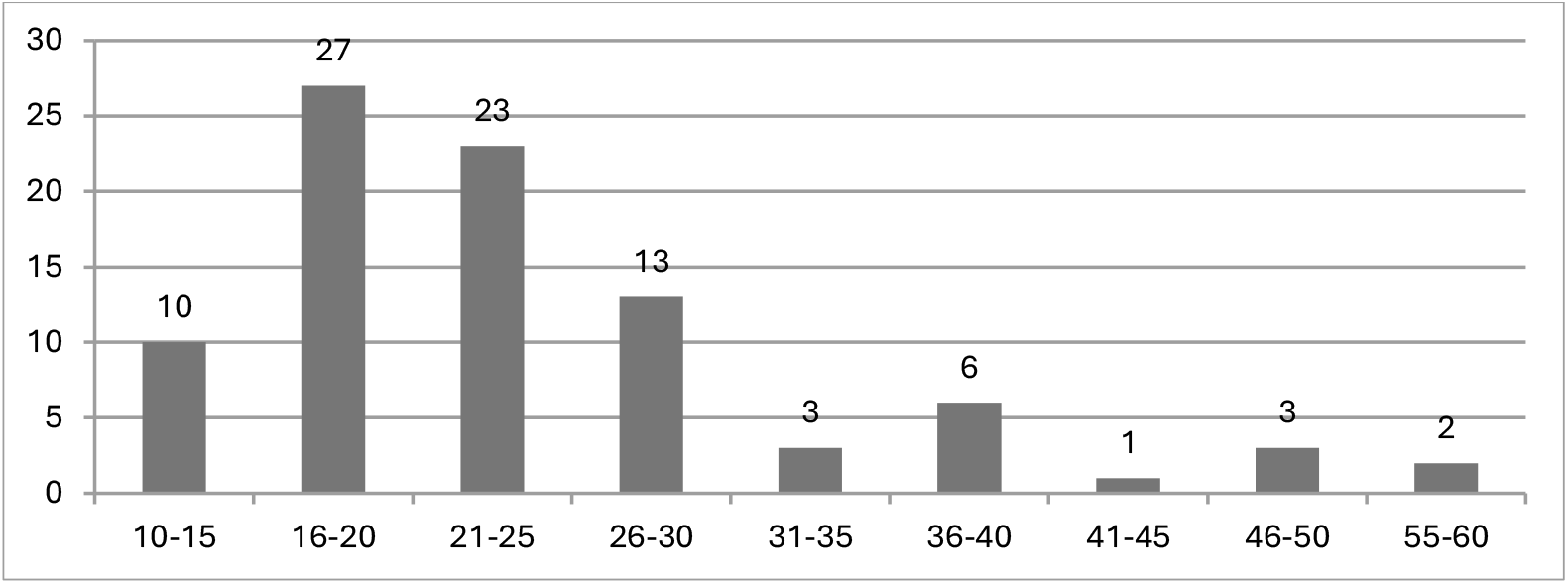
Age distribution.

Fatality rates peak in the 16-25 range, highlighting heightened vulnerability in this cohort. While event frequency decreases with age, there is a smaller peak in fatality among those aged 66-70, possibly due to age-related health changes. (Fig 2)

**Figure 2:**
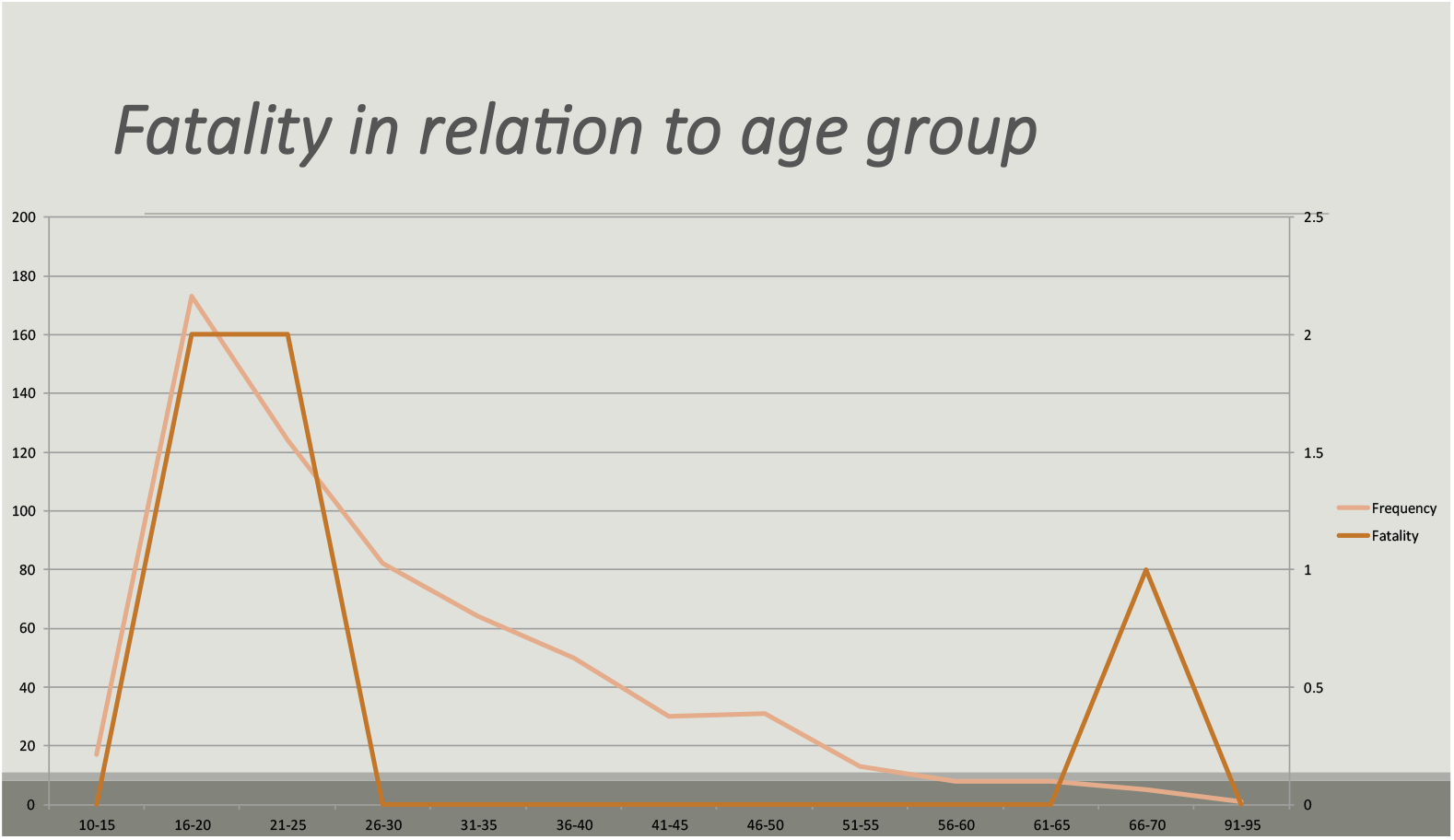
Fatality rate in relation to age.

Pesticide poisoning cases were almost evenly distributed between organophosphorus compounds (51%) and non-organophosphorus compounds (49%). Among these, organophosphorus compounds represented 8.8%. Among non-organophosphorus poisonings, a significant majority (74%) were due to “Other Pesticide/Miticide” exposures. Carbamates accounted for 7% of non-organophosphorus cases, indicating a moderate contribution.

The remainder of non-organophosphorus cases included herbicides (5%), rodenticides (2%), and other unspecified toxic agents (12%). (Fig 3 & 4)

**Figure 3:**
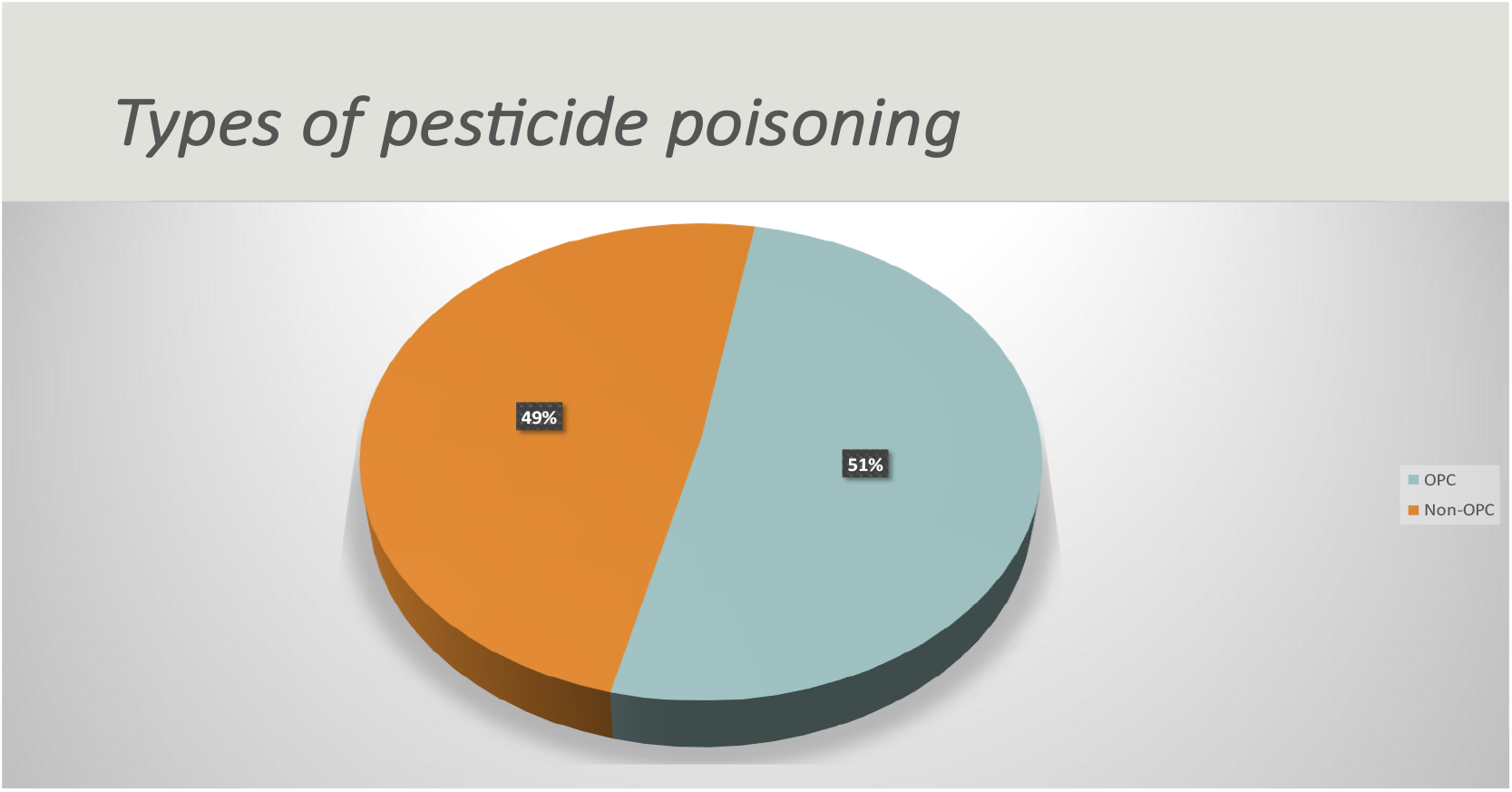
Types of Pesticide poisoning.

**Figure 3:**
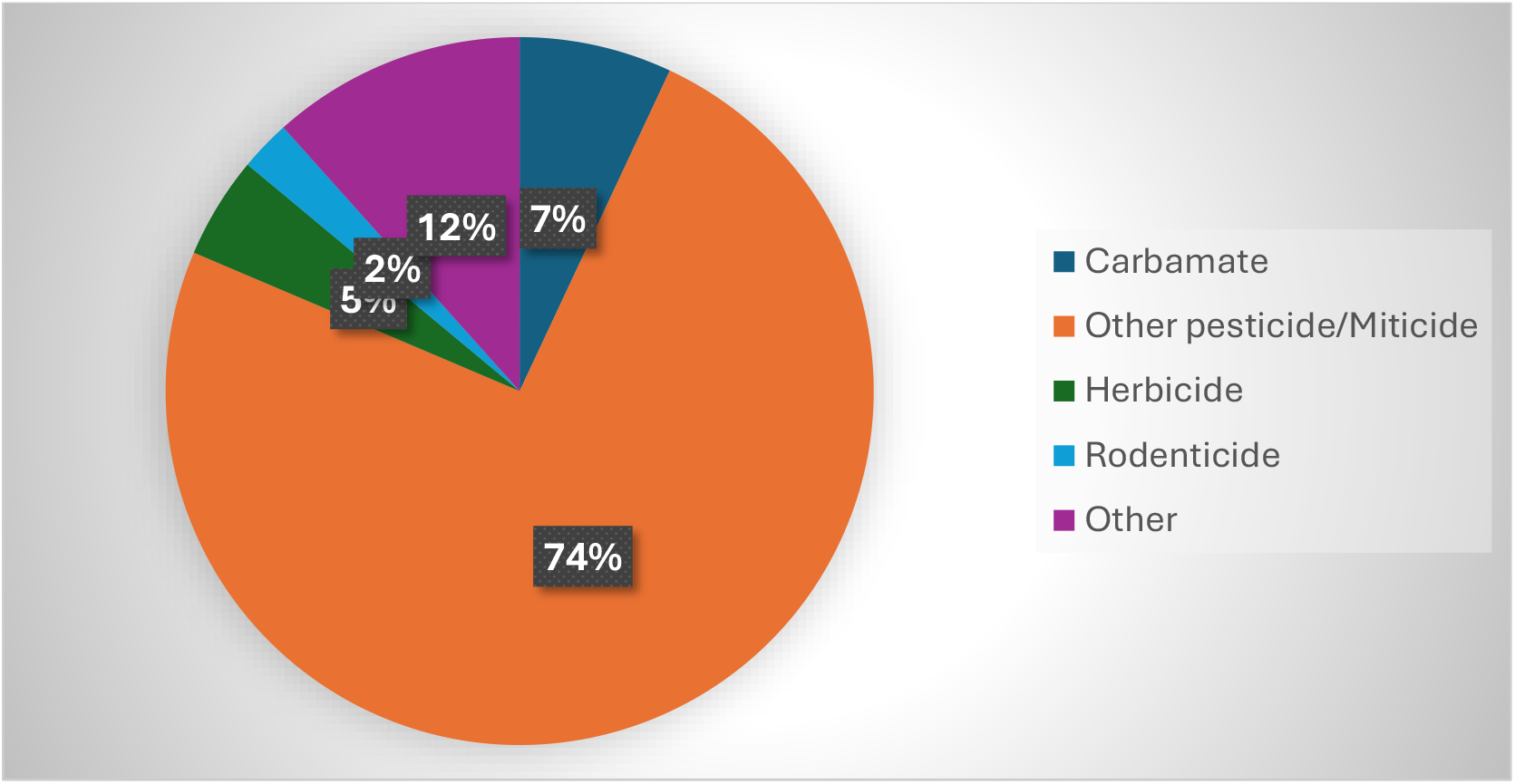
Distribution of non-OPC poisoning cases.

Commuter poisoning (31.2%) emerged as the leading type of non-pesticide poisoning, predominantly linked to criminal activities like robberies. Benzodiazepine poisoning accounted for 13.6% of cases, reflecting its easy accessibility and potential for misuse. Snake bites were reported in 7.5% of cases, underscoring significant vulnerability in rural and peri-urban areas. Household chemicals such as Harpic (7.5%), hydrogen peroxide (2.3%), and Savlon (0.8%) posed notable household risks. Medication-related poisonings included paracetamol (2.3%), sedatives (1.4%), and drug overdoses (1.1%), while illicit drug poisoning involving Yaba accounted for 0.3%. Less frequent cases included kerosene ingestion (0.8%), hair oil (0.9%), copper sulphate (0.5%), puffer fish poisoning (0.3%), and bee stings (0.2%). (Supplementary:1)

Organophosphates (OP) and other pesticides had the highest fatalities, with two deaths each, corresponding to a mortality rate of 0.3%. Categories like benzodiazepines, commuter poisoning, and drug overdose showed 100% survival rates. The highest fatality risk was linked to kerosene ingestion, which had a 20% mortality rate, despite the low number of cases.

Suicidal poisoning accounted for most cases (57.4%), followed by stupefying (32.0%) and accidental poisoning (9.8%), while homicidal (0.6%) and habitual poisoning (0.2%) were rare. The overall mortality rate for suicidal poisoning was 1.4%, with family disharmony being the most frequently reported underlying factor (indicating a 98.8% survival rate). Although family disharmony had an odds ratio (OR) of 2.18 for mortality, the wide confidence interval (CI: 0.36-13.14) and a non-significant p-value (0.395) indicate statistical uncertainty. Other motives for suicidal poisoning, such as failure to pass exams and economic loss, did not result in any fatalities. Homicidal, accidental, and stupefying poisoning cases demonstrated a 100% survival rate, highlighting the relatively low lethality of non-suicidal cases. (Supplementary: 2)

Logistic regression analysis of poisoning-related mortality suggests higher fatality rates among females (1.1%) compared to males (0.6%) (OR = 1.86, p = 0.500) and in rural areas (1.8%) compared to urban settings (0.5%) (OR = 3.75, p = 0.150). Age 26 and above showed a higher mortality rate (1.3%, OR = 3.75, p = 0.23). Those with illiteracy (2.5%, OR = 3.26) and primary education (1.4%) had the highest death rates, while individuals with higher education levels reported no fatalities. Students and homemakers both had the highest occupational mortality rate (1.2%). Although trends suggest that residing in rural areas, having lower education levels, and being female may increase risk, statistical significance was not established, indicating a need for further research. (Supplementary:3 & 4)

Logistic regression analysis of physiological parameters in poisoning-related mortality underlines several significant risk factors. A Glasgow Coma Score (GCS) of less than 9 was associated with the highest mortality (60.0%) and a markedly elevated odds ratio (OR = 80.59, p < 0.001), indicating a strong correlation with fatal outcomes. Hypotension also significantly increased mortality risk, particularly with systolic blood pressure below 80 mmHg (mortality rate of 15.4%, OR = 34.85, p < 0.001) and diastolic blood pressure below 60 mmHg (6.1% mortality, OR = 11.96, p = 0.008). Tachycardia (heart rate above 100 bpm) was linked to a 6.2% mortality rate (OR = 12.62, p = 0.032). Pupillary constriction correlated with a mortality rate of 4.1% (OR = 6.68, p = 0.040), whereas normal and dilated pupils showed no significant association with mortality. These findings underscore the importance of early monitoring and aggressive management of coma severity, hypotension, tachycardia, and pupillary abnormalities in critically poisoned patients. (Supplementary: 5)

A comparison between organophosphorus compounds (OPC) and non-organophosphorus poisoning revealed significant physiological differences. Non-OPC cases showed higher rates of hypotension (systolic blood pressure <80 mmHg: 1.7% vs. 0.6%, p = 0.027; diastolic blood pressure <60 mmHg: 4.1% vs. 1.6%, p <(0.001)and neurological impairment (GCS <11: 4.5% vs. 1.2%, p = 0.011). Tachycardia was more common in non-OPC cases (1.6% vs. 1.1%, p < 0.001.

Pupillary constriction was more prevalent in OPC poisoning (4.7% vs. 4.0%), whereas pupil dilation was more frequent in non-OPC cases (8.4% vs. 2.9%, p < 0.001). These findings suggest that non-OPC poisoning is associated with more severe hypotension, neurological impairment, and pupil dilation, whereas OPC poisoning primarily causes pupillary constriction. (Supplementary:6)

## Discussion

The study reveals critical challenges in managing acute poisoning in Bangladesh, highlighting commuter poisoning, pesticide exposure, and snake bites as primary public health threats. The high incidence of commuter poisoning underscores the urgent need for improved security measures, community vigilance, and public safety education to minimize intentional harm and robberies.

Pesticide poisoning, especially from organophosphorus compounds, remains prevalent and reflects ongoing challenges in agricultural practices and chemical handling. ^7^ Promoting safer storage practices, appropriate labelling, and educating farming communities about pesticide risks are essential steps in prevention.

Moreover, strengthening surveillance and prompt clinical interventions can mitigate adverse outcomes in pesticide poisonings.

Our study identified that organophosphorus compounds were responsible for 8.8% of poisoning cases, while other pesticides accounted for 10.5%. This aligns with regional data indicating that organophosphates are commonly implicated in poisoning incidents due to their widespread use in agriculture.^7^ It underscores the importance of stringent regulatory frameworks and the promotion of safe agricultural practices to mitigate the risks associated with pesticide use.

Similar trends have been noted globally, highlighting the critical importance of implementing robust regulatory frameworks, as emphasized by studies such as those conducted by Eddleston et al. (2008), Gunnell et al. (2007), and Mishara (2007). ^2,3,4^ Moreover, strengthening surveillance and prompt clinical interventions, as recommended by WHO guidelines (WHO, 2019), can mitigate adverse outcomes in pesticide poisonings.^1^

The prevalence of snake bites highlights the need for enhanced community education programs about preventive measures and appropriate immediate responses following snake bites. Ensuring the widespread availability of antivenom and training healthcare providers on proper snakebite management is crucial.

Suicidal poisoning is the most prevalent and carries the highest risk, often linked to psychosocial stressors such as family conflicts, academic struggles, and financial distress. In contrast, homicidal and accidental poisoning cases are less common and typically result in better survival rates.

Demographic, socioeconomic, and physiological factors significantly influence poisoning-related mortality. Although females may face a slightly higher risk than males, this difference is not statistically significant. Religious background and marital status do not appear to affect survival rates. Environmental factors are more telling; rural residents have a higher mortality risk compared to urban residents, likely due to limited healthcare access and exposure to toxic substances. Age also plays a role, with older individuals typically at greater risk due to underlying health conditions.

Education and occupation matter as well. Lower educational attainment is linked to higher mortality rates, indicating that awareness of toxic substances can be protective. Students and housewives may also be more vulnerable due to differences in exposure and response to treatment. These findings highlight the need for targeted interventions, including improved healthcare access in rural areas and educational programs to enhance awareness and prevention of poisoning.

Physiological parameters are crucial for predicting mortality related to poisoning. The Glasgow Coma Scale (GCS) is a key indicator of neurological status, where lower scores are linked to a significantly higher risk of death. Similarly, blood pressure abnormalities, particularly hypotension, indicate systemic instability and poor prognosis, reinforcing the importance of early hemodynamic support. Heart rate abnormalities also contribute to poisoning outcomes.

Organophosphate poisoning shows unique physiological patterns, featuring symptoms like pupillary constriction, whereas non-organophosphate poisonings present a wider range of disturbances.

The findings emphasize the importance of early clinical assessment, continuous vital sign monitoring, and aggressive supportive care in poisoning management. Targeted interventions such as timely resuscitation and toxin-specific antidotes are crucial for reducing mortality in severe cases. Further research is needed to enhance predictive models and improve triage strategies for better patient outcomes.

## Conclusion

This study highlights the critical gaps in managing acute poisoning cases in Bangladesh, necessitating immediate action through structured clinical protocols, improved healthcare provider training, and targeted community-based interventions. Effective public awareness campaigns, regulatory reinforcement, and resource allocation for rapid clinical response can significantly mitigate poisoning-related morbidity and mortality. Implementing these comprehensive strategies is essential for enhancing healthcare system preparedness and ultimately improving patient outcomes.

## Data Availability

All data are available in the manuscript and supplementary data

