## Supplementary files for "Two-Month Study on Acute Poisoning Cases in a Tertiary Care Hospital in Bangladesh"

Supplementary 1

| Variable | Total/Missing data | Of All | | Survivors | | Deaths | | Mortality |
|---|---|---|---|---|---|---|---|---|
| | 611/39 | N | % | N | % | N | % | |
| OP | | 54 | 8.8% | 52 | 96.3% | 2 | 3.7% | 0.3% |
| Carbamate | | 5 | 0.8% | 5 | 100% | 0 | 0 | 0 |
| Other pesticide | | 64 | 10.5% | 62 | 96.9% | 2 | 3.1% | 0.3% |
| Benzodiazepines | | 83 | 13.6% | 83 | 100% | 0 | 0 | 0 |
| Other sedatives | | 4 | 0.7% | 4 | 100% | 0 | 0 | 0 |
| Kerosine | | 6 | 0.8% | 4 | 80% | 1 | 20% | 0.2% |
| Snake bite | | 46 | 7.5% | 46 | 100% | 0 | 0 | 0 |
| Copper Sulphate | | 3 | 0.5% | 3 | 100% | 0 | 0 | 0 |
| Potka fish | | 2 | 0.3% | 2 | 100% | 0 | 0 | 0 |
| Others | | 345 | 56.1% | 343 | 100% | 0 | 0 | 0 |
| Commuter poisoning | | 203 | 31.2% | 203 | 100% | 0 | 0 | 0 |
| Harpic poisoning | | 49 | 7.5% | 49 | 100% | 0 | 0 | 0 |
| Hydrogen Peroxide | | 15 | 2.3% | 15 | 100% | 0 | 0 | 0 |
| Paracetamol | | 15 | 2.3% | 15 | 100% | 0 | 0 | 0 |
| Multiple | | 9 | 1.4% | 9 | 100% | 0 | 0 | 0 |
| Hair Oil | | 6 | 0.9% | 6 | 100% | 0 | 0 | 0 |
| Drug Overdose | | 7 | 1.1% | 7 | 100% | 0 | 0 | 0 |
| Savlon | | 5 | 0.8% | 5 | 100% | 0 | 0 | 0 |
| Bee bite | | 1 | 0.2% | 1 | 100% | 0 | 0 | 0 |
| Yaba | | 2 | 0.3% | 2 | 100% | 0 | 0 | 0 |
| Unknown | | 40 | 6.2% | 40 | 100% | 0 | 0 | 0 |

Supplementary 2

### *Cause and Motive of poisoning*

| Variable | Total (%)/Missing data | Category | Proportion of deaths | Of All | | Survivors | | Deaths | | Odds ratio | 95% CI | P value |
|---|---|---|---|---|---|---|---|---|---|---|---|---|
| Cause | 603/47 | | | N | % | N | % | N | % | | | |
| Suicidal | 368 (57.4%) | | 1.4% | 5 | 0.8% | 343 | 98.6% | 5 | 1.4% | NA | | |
| | | Family Disharmony | 1.2% | 3 | 0.9% | 247 | 98.8% | 3 | 1.2% | 2.18* | 0.36-13.14 | 0.395 |
| | | Fail to Pass Exam | 0 | 0 | 0 | 2 | 100% | 0 | 0 | NA | | |
| | | Economic Loss | 0 | 0 | 0 | 1 | 100% | 0 | 0 | NA | | |
| | | Others | 1.1% | 1 | 0.3% | 88 | 98.9% | 1 | 1.1% | 1.47* | 0.16-13.32 | 0.73 |
| Homicidal | 4 (0.6%) | | 0 | 0 | 0 | 4 | 100% | 0 | 0 | NA | | |
| Accidental | 63 (9.8%) | | 0 | 0 | 0 | 61 | 100% | 0 | 0 | NA | | |
| Stupefying | 205 (32.0%) | | 0 | 0 | 0 | 189 | 100% | 0 | 0 | NA | | |
| Others (habitual) | 1 (0.2%) | | 0 | 0 | 0 | 1 | 100% | 0 | 0 | NA | | |

As all deaths were in suicidal group, OR can not be calculated.

Supplementary 3

### *Logistic regression analysis and Odds Ratio (OR) of gender, religion, marital statu and age in causing death of patients*

| Variable | Total/Missing data | Category | Proportion of deaths | Of All | | Survivors | | Deaths | | Odds ratio | 95% CI | P value |
|---|---|---|---|---|---|---|---|---|---|---|---|---|
| | | | | N | % | N | % | N | % | | | |
| Gender | 609/41 | Male | 0.6% | 2 | 0.3% | 334 | 99.4% | 2 | 0.6% | 1 | | |
| | | Female | 1.1% | 3 | 0.5% | 270 | 98.9% | 3 | 1.1% | 1.86 | 0.31-11.18 | 0.500 |
| Religion | 575/75 | Islam | 0.9% | 5 | 0.9% | 567 | 99.1% | 5 | 0.9% | NA | | |
| | | Hindu | 0 | 0 | 0 | 3 | 100% | 0 | 0 | NA | | |
| | | Buddhist | 0 | 0 | 0 | 0 | 0 | 0 | 0 | NA | | |
| Marital Status | 573/77 2 divorced case excluded | Married | 0.8% | 3 | 0.5% | 362 | 99.2% | 3 | 0.8% | 1 | | |
| | | Unmarried | 1.0% | 2 | 0.3% | 206 | 99.0% | 2 | 1.0% | 1.17 | 0.19-7.06 | 0.862 |
| Setting | 586/64 | Rural | 1.8% | 3 | 0.5% | 166 | 98.2% | 3 | 1.8% | 3.75 | 0.62-22.65 | 0.150 |
| | | Urban | 0.5% | 2 | 0.3% | 415 | 99.5% | 2 | 0.5% | 1 | | |
| Age | 606/44 | <16 years | 0 | 0 | 0 | 17 | 100% | 0 | 0 | NA | | |
| | | ≥16 years | 0.8% | 5 | 0.8% | 584 | 99.2% | 5 | 0.8% | NA | | |
| | | <26 years | 1.3% | 4 | 0.7% | 310 | 98.7% | 4 | 1.3% | 3.75 | 0.41-33.79 | 0.23 |

Supplementary 4

### *Logistic regression analysis and Odds Ratio (OR) of education and occupation in c[illegible] death of patients*

| Variable | Total/Missing data | Category | Proportion of deaths | Of All | | Survivors | | Deaths | | Odds ratio | 95% CI | P value |
|---|---|---|---|---|---|---|---|---|---|---|---|---|
| | | | | N | % | N | % | N | % | | | |
| Education | 554/96 | Illiterate | 2.5% | 1 | 0.2% | 39 | 97.5% | 1 | 2.5% | 3.26 | 0.36-29.96 | 0.295 |
| | | Primary | 1.4% | 2 | 0.4% | 137 | 98.6% | 2 | 1.4% | 2.01 | 0.33-12.12 | 0.449 |
| | | High School | 0.4% | 1 | 0.2% | 247 | 99.6% | 1 | 0.4% | 0.31 | 0.03-2.75 | 0.291 |
| | | Intermediate | 1.1% | 1 | 0.2% | 94 | 98.9% | 1 | 1.1% | 1.21 | 0.13-10.94 | 0.87 |
| | | Graduate and above | 0 | 0 | 0 | 32 | 100% | 0 | 0 | NA | | |
| | | Above Graduate | 0 | 0 | 0 | 1 | 100% | 0 | 0 | NA | | |
| Occupation | 558/92 | Service | 0 | 0 | 0 | 49 | 100% | 0 | 0% | NA | | |
| | | Farmer | 0 | 0 | 0 | 30 | 100% | 0 | 0 | NA | | |
| | | Student | 1.2% | 2 | 0.4% | 160 | 98.8% | 2 | 1.2% | 1.64 | 0.27-9.89 | 0.591 |
| | | Housewife | 1.2% | 2 | 0.4% | 169 | 98.8% | 2 | 1.2% | 1.52 | 0.25-9.14 | 0.651 |
| | | Businessman | 0 | 0 | 0 | 53 | 100% | 0 | 0 | NA | | |
| | | Others | 1.1% | 1 | 0.2% | 92 | 98.9% | 1 | 1.1% | 1.25 | 0.13-11.34 | 0.841 |

Supplementary 5

*Logistic regression analysis and Odds Ratio (OR) of GCS, blood pressure, heart rate and condition of pupil in causing death of patients.*

| Variable | Total/Missing data | Category | Proportion of deaths | Of All | | Survivors | | Deaths | | Odds ratio | 95% CI | P value |
|---|---|---|---|---|---|---|---|---|---|---|---|---|
| | | | | N | % | N | % | N | % | | | |
| | Total data 650 | | | | | | | Total death 5 | | | | |
| Glasgow coma score | 607/43 | <9 | 60.0% | 3 | 0.5% | 11 | 78.6% | 3 | 21.4% | 80.59 | 12.22-531.32 | <0.001 |
| Blood pressure | 591/59 | Systolic <80 | 15.4% | 2 | 0.3% | 11 | 84.6% | 2 | 15.4% | 34.85 | 5.29-229.76 | <0.001 |
| | | Diastolic <60 | 6.1% | 2 | 0.3% | 31 | 93.9% | 2 | 6.1% | 11.96 | 1.92-74.19 | 0.008 |
| Heart rate | 587/63 | >100 | 6.2% | 1 | 0.2% | 15 | 93.8% | 1 | 6.2% | 12.62 | 1.24-128.50 | 0.032 |
| | | <60 | 0 | 0 | 0 | 4 | 100% | 0 | 0% | NA | | |
| Pupil | 585/65 | Normal | 0.6% | 3 | 0.5% | 471 | 99.4% | 3 | 0.6% | 1 | | |
| | | Dilated | 0 | 0 | 0 | 62 | 100% | 0 | 0% | NA | | |
| | | Constricted | 4.1% | 2 | 0.3% | 47 | 96.9% | 2 | 0.3% | 6.68 | 1.09-40.99 | 0.040 |

Supplementary 6

### *Comparison of vital signs and other features in between OPC and non-OPC poisoning group*

| | | OPC (n =62) | of Total | Non-OPC (n = 588) | of Total | p-value | Note |
|---|---|---|---|---|---|---|---|
| **Systolic BP** | <80 mmHg | 4 | 0.6% | 11 | 1.7% | 0.027 | |
| **Diastolic BP** | <60 mmHg | 10 | 1.6% | 26 | 4.1% | <0.001 | |
| **HR** | < 60 beats/min | 0 | 0.0% | 4 | 0.6% | 0.521 | |
| | >100 beats/min | 7 | 1.1% | 10 | 1.6% | <0.001 | |
| **GCS** | <11 | 9 | 1.2% | 29 | 4.5% | .011 | In comparison to >11 |
| | 11 to 14 | 19 | 2.9% | 155 | 23.8% | 0.26 | In comparison to GCS 15 |
| **Pupil** | Constricted | 29 | 4.7% | 25 | 4.0% | <0.001 | In comparison to normal pupil |
| | Dilated | 18 | 2.9% | 52 | 8.4% | <0.001 | In comparison to normal pupil |
| | Normal | 15 | 2.4% | 483 | 77.7% | <0.001 | In comparison to constricted |
